# Performance of a foundation model to generate thousands of predicted outcomes across multiple use cases

**DOI:** 10.64898/2026.10.28.26366723

**Authors:** Matteo Mischiati, Kyriakos Vattis, Richard Gliklich, Michelle Leavy, Costas Boussios, Jigar Bandaria

## Abstract

**Background:** Predictive artificial intelligence (AI) models hold great promise for medical care. While many types of models have proliferated across healthcare and the life sciences, systematic, rapid generation of multi-condition predictions of diagnoses, future medical events and future clinical outcomes from a single foundation-model architecture remains rare. PhenOM® is a generative AI foundation model trained and validated on longitudinal medical-event data from more than 400 million patient records in the OM1 Real World Data Cloud (RWDC) and designed to complete predictive tasks for a broad array of medical diagnoses, events and clinical outcomes (collectively ‘outcomes’) at varying time intervals, currently up to five (5) years. This paper describes the implementation of PhenOM to complete a vast number of predictive tasks across clinical domains and outcomes.

**Methods:** PhenOM was evaluated across four outcome categories (Future Risk, Screening, Care Opportunity, and Treatment Effect) in multiple domains spanning chronic disease, acute medical events, healthcare utilization, and rare and frequently undiagnosed conditions. Performance was assessed using area under the receiver operating characteristic curve (AUROC). Outcomes were defined within an eligible-population framework using standardized medical code sets (ICD-10, SNOMED, CPT, NDC, LOINC) unless outcomes could not be specified with an existing code. All benchmarks reported were derived from data excluded from training.

**Results:** As of this publication, the single PhenOM model has been used to complete and validate thousands of predictive tasks covering more than 1,770 outcomes spanning most organ systems and clinical specialties, with new predictive tasks being validated each day. Overall, 80% of predictive tasks achieved AUROC of at least 0.70 and 30% achieved AUROC of at least 0.80. We present a sample of 177 of these tasks as examples that show the breadth of tasks the model can complete. Performance varied systematically by outcome and clinical domain.

**Conclusions:** PhenOM demonstrates the ability to complete thousands of predictive tasks across clinical domains and outcomes at the individual patient level, without additional training. Outcomes include screening for undiagnosed conditions, predicting future conditions, medical events, effects of treatments, and assessing care opportunities. These can be ascertained at the individual patient and population levels. These findings support applications in individual and population health management, risk management, drug development, and real-world evidence generation, and establish a transparent, reproducible performance reference for a large-scale real-world predictive AI platform.

## Introduction

Real-world data from EHRs, claims, and registries have increasingly been used to train and validate clinical prediction models. Machine-learning approaches, including gradient-boosted trees, recurrent neural networks, and transformer-based architectures, have shown predictive performance across a wide range of conditions.^1^ Yet, most published models address a single outcome within a specific disease context, and systematic comparisons across outcome types, clinical domains, and patient populations remain rare. Value-based care models ultimately rely on effective prognostication. Health systems, providers, payers, and life-sciences organizations increasingly need tools that can generate risk estimates for many outcomes from a common patient-data infrastructure, rather than building and maintaining a separate model for each question.

Foundation models, which are single, typically large models trained on broad data and then adapted to many downstream tasks, have emerged as a promising approach for flexible clinical prediction. Unlike task-specific models, they encode a generalizable representation of patient health trajectories, allowing many outcomes to be predicted without separate model development for each target. The concept of a general-purpose foundation model trained on longitudinal EHR data has been explored through several architectures. Early deep-learning work showed that clinical events represented in a standardized (FHIR) format could predict multiple outcomes simultaneously without site-specific data harmonization.^2^ Subsequent approaches, including BEHRT^3^ and Med-BERT,^4^ extended this paradigm using masked pretraining, and more recent generative medical-event models have shown that predictive performance improves according to consistent scaling laws as model and data size increase.^5^ A recurring finding is that models trained on large, diverse data can perform as well as, or better than, task-specific fine-tuned models on held-out outcomes, supporting the foundation-model hypothesis for clinical data.

Most academic foundation models, however, are trained on data from a single health system or a small number of systems, which limits their representativeness and generalizability.^6^ A recent multimodal, temporal EHR foundation model (APOLLO) illustrates both the potential of these models and this constraint. APOLLO was trained on 25 billion medical events from 7.2 million patients across a single multi-institutional US health system over three decades. It produced whole-patient (“virtual patient”) representations and was evaluated across more than 300 prognostic and retrieval tasks, but its broad applicability is likely limited by being trained on a single academic health system’s population.^7^

PhenOM uses data from a patient’s medical-event history, including diagnoses, medications, laboratory results, procedures, mortality, and other data streams. Four outcome categories are supported: Future Risk (new conditions or events), Screening (undiagnosed conditions), Care Opportunity (gaps in care), and Treatment Effect (the predicted effect of a treatment).

The purpose of this paper is to describe the use of PhenOM to complete predictive tasks for outcomes in all four categories at scale from real-world data. PhenOM has generated thousands of predictive tasks to date. We present a review of a ten percent representative sample (177) of rigorously validated tasks in the catalog as examples that illustrate the breadth of clinical applicability, the performance achievable across patient populations, and the practical framework for defining and evaluating outcomes. New predictive tasks can be completed as additional use cases and research questions arise. Our aim is to give the research and clinical community a transparent, reproducible view of what a large-scale, real-world predictive AI platform can produce.

## Methods

### Data source: OM1 Real World Data Cloud

All analyses were conducted using non-training data from the OM1 Real-World Data Cloud™ (“RWDC”), a de-identified, multi-source, deterministically linked, fully integrated real-world healthcare data warehouse. The RWDC includes data from electronic health records (EHRs), laboratories, medical and pharmacy claims and mortality sources and is organized as de-identified patient longitudinal records. Structured data is coded in standard terminologies (ICD-10, ICD-9, SNOMED, CPT, HCPCS, NDC, RxNorm, GPI, and LOINC). Unstructured data is converted into well-defined, structured concepts. As of July 2026, the database contains healthcare data on over 400 million patients in the United States, with data collected since 2013. The average duration of patients in the database is over 10 years. By census region, the distribution of the RWDC population largely reflects the overall US population. Importantly, data are representative of U.S. medical practice in terms of community, rural, academic, and tertiary centers. The protocol describing use of the RWDC for research on patient clinical characteristics and outcomes was reviewed by the Advarra Institutional Review Board and determined to be exempt.

### PhenOM Foundation Model

PhenOM is a foundation model trained on longitudinal medical-event sequences. For each outcome–patient pair, it generates a continuous score representing the predicted likelihood of a clinical event within a defined time window, or the likelihood of a screening- or treatment-eligible status.

PhenOM is a ∼1 billion parameter GPT Transformer model with a vocabulary size of ∼280,000. PhenOM is trained on a set of 200 million patients from the OM1 RWDC, a de-identified database that combines data from EMRs and administrative sources.

Patients have an average of greater than 10 years of medical history data. The OM1 RWDC contains a total of about 400 million patients. The ∼200 million patients not used for training serves as a holdout set that is used to validate the various predictive tasks generated from the foundation model, PhenOM. For each predictive task, eligibility criteria, including demographics, comorbidities and completeness of history, for a set of patients is drawn from the holdout set to form the validation set. Given its size, the holdout set covers practically all ages and all US census regions.

A detailed description of the model architecture, training objective, and scale will be provided in a forthcoming publication on PhenOM model development.

### Outcome types and the eligible-population framework

One useful categorization of predictions is by the type of outcome each predicts, with the category reflecting how the prediction could be used in clinical practice. Alternative framing could be used for other use cases such as in risk management or population analytics. We report results herein using the following categories:

- **Future Risk**: the likelihood that a patient will experience a medical event, such as the onset of a new condition or an acute event, within a defined time window. Used for proactive risk stratification, counseling and preventive intervention.
- **Screening**: patients ranked by the likelihood of having an undiagnosed condition — those whose longitudinal record is consistent with a diagnosis not yet formally recorded. Used to prioritize candidates for diagnostic workup.
- **Care Opportunity**: patients ranked by the likelihood of being eligible for a specific treatment or therapy not yet received. Used to identify undertreated patients.
- **Treatment Effect:** the likelihood a patient will have a specific effect from a specific treatment in a specified timeframe. Used to predict the impact of a specific medication or procedure on an individual patient. (Note, these tasks estimate the risk of an outcome among patients who received the treatment; they are not estimates of causal treatment effects.)

Each predictive task is defined relative to an eligible population characterized by inclusion criteria (for example, minimum age, relevant medical history, and absence of the outcome in prior history). Outcomes are operationalized using precise definitions that leverage definitional standards (e.g., standard code sets) whenever possible. This framework supports reproducibility and enables direct comparison of performance across predictive tasks.

### Predictive tasks and validation approach

Predictive tasks are created through an iterative, repeatable process involving clinical subject-matter experts, data scientists, and product managers, allowing predictive tasks for new outcomes to be defined and the PhenOM model tested for performance. Each predictive task is assigned a stable identifier, a standardized and reproducible definition, and documented inclusion criteria. Performance was evaluated using AUROC. Throughout, an outcome is a clinical condition or event (e.g., heart failure), and a predictive task is a specific combination of outcome, eligible population, and time window; a single outcome may therefore have several predictive tasks.

## Results

### Predictive tasks completed to date and outcome category and clinical domain coverage

To date, PhenOM has generated and validated thousands of predictive tasks with 1770 included in the catalog at the time this analysis was performed. This set continues to expand as new use cases arise. Across all catalog tasks, 80% achieved AUROC of at least 0.70, and 30% achieved AUROC of at least 0.80. This paper presents a representative sample of 177 of these tasks that show the breadth of tasks the model can complete; representative examples are summarized by clinical domain in Table 1. The full set of predictive tasks completed to date is distributed across the four outcome categories and represents more than 12 clinical domains or utilization areas and discrete time periods for future risks (i.e. 1, 3, 6, 12, 24, 36, 48 or 60 months). Clinical domains represented include behavioral health, cardiovascular disease, diabetes and other metabolic conditions, frailty, gastrointestinal conditions, infectious diseases, musculoskeletal conditions, neurological and balance disorders, oncology, women’s health, and renal disorders. In addition, major surgical procedures across all health conditions are included. Healthcare utilization (e.g., hospitalization and emergency-department use) and mortality outcomes are also available.

**Table 1.** Examples of PhenOM predictive tasks completed to date.

| Clinical domain | Example validated outcomes (outcome type) | AUROC range of listed examples |
| --- | --- | --- |
| Cardiovascular & circulatory | Heart failure, atrial fibrillation, myocardial infarction, ischemic stroke (Future Risk); aortic valve stenosis, peripheral arterial disease (Screening); PCSK9 inhibitors, direct oral anticoagulants in atrial fibrillation (Care Opportunity); stroke after CABG, new-onset atrial fibrillation after CABG (Treatment Effect) | 0.69–0.87 |
| Metabolic & endocrine | Type 2 diabetes, type 1 diabetes, hypoglycemia in diabetes (Future Risk); primary aldosteronism, Cushing's syndrome (Screening); GLP-1 receptor agonists for diabetes, GLP-1 receptor agonists for weight loss (Care Opportunity) | 0.73–0.86 |
| Renal | Chronic kidney disease, ESRD in type 2 diabetes, hyperkalemia in CKD (Future Risk); undiagnosed CKD, renal artery stenosis (Screening); acute kidney injury after CABG (Treatment Effect) | 0.65–0.87 |
| Neurological & cognitive | Alzheimer's disease, dementia (Future Risk); Parkinson's disease, epilepsy, multiple sclerosis (Screening); amyloid-targeting therapies in MCI or Alzheimer's, migraine treatment (Care Opportunity); post-operative delirium after SHFFT (Treatment Effect) | 0.75–0.92 |
| Behavioral health | Substance use overdose, suicide attempt, major depression onset (age $\geq 65$) (Future Risk); major depression, bipolar disorder, substance use disorder, generalized anxiety disorder (Screening); new chronic opioid use after spinal fusion (Treatment Effect) | 0.67–0.83 |
| Oncology | Lung cancer, lung cancer in COPD, breast cancer, all-cause hospitalization in cancer (Future Risk); multiple myeloma, prostate cancer, colorectal cancer, venous thromboembolism in cancer (Screening) | 0.72–0.84 |
| Respiratory | Spontaneous pneumothorax in COPD, respiratory failure in COPD (Future Risk); idiopathic pulmonary fibrosis, pulmonary arterial hypertension, COPD, asthma (Screening) | 0.73–0.91 |
| Musculoskeletal & surgical | Total knee replacement, lumbar spine surgery, fragility fracture in osteoporosis (Future Risk); rheumatoid arthritis, osteoarthritis (Screening); benefit after total knee replacement, VTE after total hip replacement, periprosthetic joint infection after TKR (Treatment Effect) | 0.71–0.82 |
| Rare & underdiagnosed conditions | Portal vein thrombosis, Sjögren disease, ALS, systemic sclerosis, ATTR amyloidosis in heart failure, Fabry disease (Screening) | 0.63–0.84 |
| Women's health | Osteoporosis (females $\geq 45$), polycystic ovary syndrome, endometriosis (Screening) | 0.68–0.77 |
| Gastrointestinal & hepatic | Chronic pancreatitis, MASH, Barrett's esophagus (Screening); surgical site infection after major bowel procedures, anastomotic leak (Treatment Effect) | 0.63–0.82 |
| Healthcare utilization & mortality | All-cause mortality, all-cause hospitalization, emergency department visit (Future Risk); 30-day all-cause readmission after SHFFT, TKR, THR, or CABG (Treatment Effect) | 0.61–0.87 |

*Examples are representative, not exhaustive, and were selected to illustrate each of the four outcome types where available; AUROC ranges are the minimum and maximum AUROC of the examples listed in each row. ALS, amyotrophic lateral sclerosis; ATTR, transthyretin amyloidosis; CABG, coronary artery bypass grafting; CKD, chronic kidney disease; COPD, chronic obstructive pulmonary disease; ESRD, end-stage renal disease; GLP-1, glucagon-like peptide-1; MASH, metabolic dysfunction-associated steatohepatitis; MCI, mild cognitive impairment; PCSK9, proprotein convertase subtilisin/kexin type 9; SHFFT, surgical hip or femur fracture treatment; THR, total hip replacement; TKR, total knee replacement; VTE, venous thromboembolism.*

### Overall performance by outcome type

Figure 1 shows the distribution of AUROC for the 149 Future Risk (n = 94) and Screening (n = 55) tasks in the 177-task sample; the remaining tasks were Care Opportunity (n = 6) and Treatment Effect (n = 22). AUROC values for Future Risk outcome predictions ranged from 0.62 to 0.92 (median 0.76; IQR 0.70–0.80), while Screening outcome predictions showed AUROC values from 0.62 to 0.91 (median 0.77; IQR 0.73–0.82). Predictions for outcomes categorized as Future Risk or Screening were randomly selected in the ratio they are currently available as validated predictions. Care Opportunity omitted since outcomes are more heterogeneous. Treatment Effect outcome predictions ranged from 0.61 to 0.85 (median 0.69; IQR 0.66–0.72); these are peri-procedural complications within surgical episode bundles or medication adverse events and are summarized separately below.

**Figure 1.**
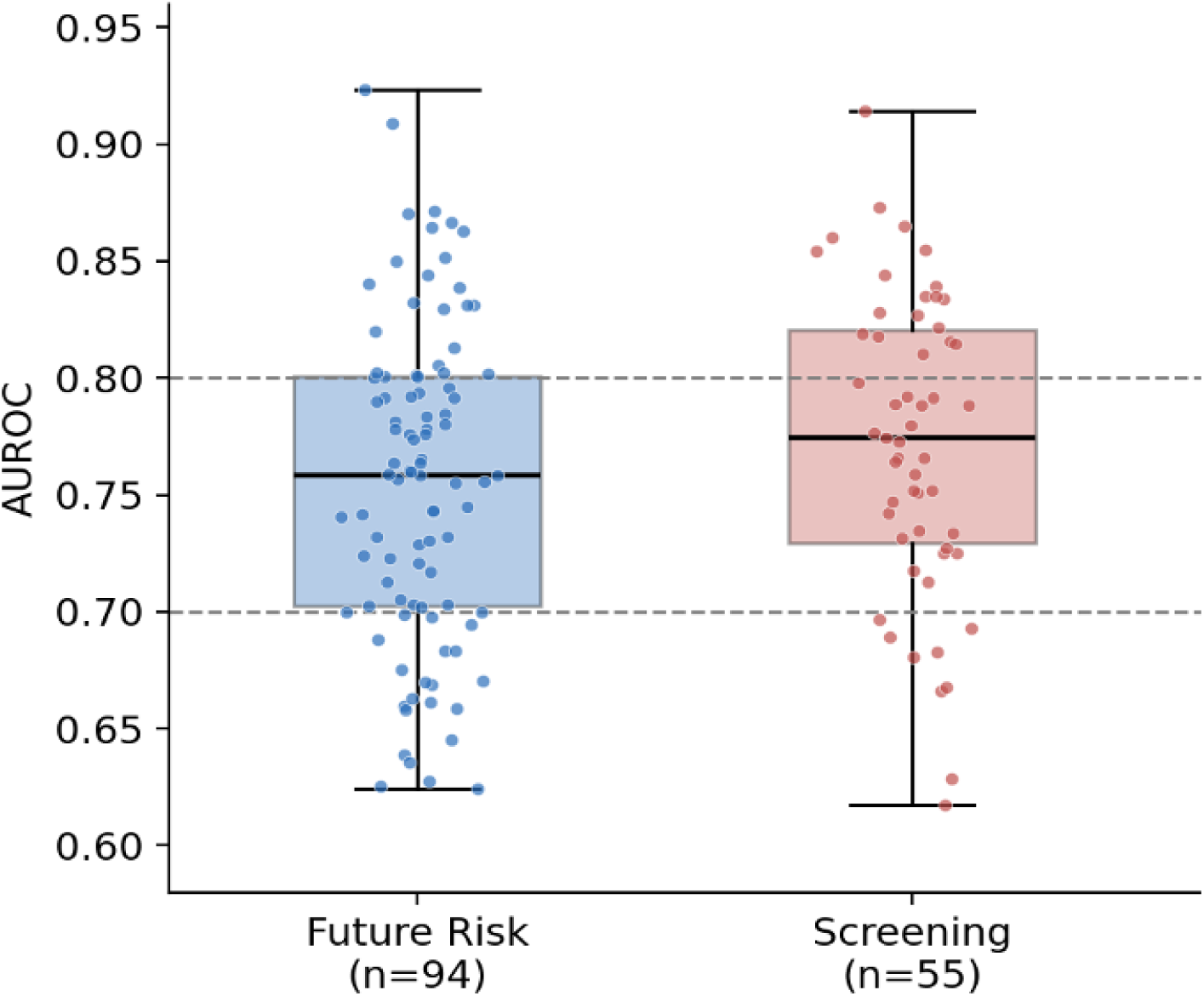
Distribution of AUROC by outcome type for Future Risk and Screening tasks in the sample.

### Performance across clinical domains

To illustrate performance heterogeneity across clinical areas, we describe representative outcomes below.

Table 2 presents the three high-performing outcomes within each outcome type. Where several cohort variants or closely related definitions of the same outcome were available, only the highest-performing variant is shown.

**Table 2.** High-performing outcomes by outcome type (three per type)

| Type | Outcome | Brief description | Cohort* | Timeframe | AUROC |
| --- | --- | --- | --- | --- | --- |
| Future Risk | Alzheimer's disease | New diagnosis of Alzheimer's disease | General population | 1 year | 0.92 |
| Future Risk | All-cause mortality | Death from any cause | General population | 1 year | 0.87 |
| Future Risk | End-stage renal disease | Progression to end-stage renal disease | Diabetes type 2 | 1 year | 0.87 |
| Screening | Idiopathic pulmonary fibrosis | IPF present but not yet diagnosed | General population | Not applicable | 0.91 |
| Screening | Pulmonary arterial hypertension | PAH present but not yet diagnosed | General population | Not applicable | 0.89 |
| Screening | Aortic valve stenosis | Aortic valve stenosis present but not yet diagnosed | General population | Not applicable | 0.87 |
| Care Opportunity | Amyloid-targeting therapies | Receipt of an amyloid-targeting therapy | MCI or Alzheimer's | Not applicable | 0.88 |
| Care Opportunity | GLP-1 receptor agonists for diabetes | Receipt of a GLP-1 receptor agonist for diabetes | General population | Not applicable | 0.86 |
| Care Opportunity | PCSK9 inhibitors | Receipt of a PCSK9 inhibitor | General population | Not applicable | 0.83 |
| Treatment Effect | Delirium | Delirium following surgical hip or femur fracture treatment | Surgical hip or femur fracture treatment | 1 month | 0.85 |
| Treatment Effect | All-cause readmission | Readmission for any cause following surgical hip or femur fracture treatment | Surgical hip or femur fracture treatment | 1 month | 0.79 |
| Treatment Effect | Venous thromboembolism | VTE following total hip replacement | Total hip replacement | 1 month | 0.75 |

*The three highest AUROC outcomes within each outcome type are shown. Where several cohort variants or closely related definitions of the same outcome were available (e.g., Alzheimer's disease and all-cause dementia; pulmonary arterial hypertension and pulmonary hypertension), only the highest-performing variant is shown. Screening outcomes estimate the likelihood that a condition is present but not yet diagnosed at the time of prediction; a prospective outcome timeframe therefore does not apply. GLP-1, glucagon-like peptide-1; IPF, idiopathic pulmonary fibrosis; MCI, mild cognitive impairment; PAH, pulmonary arterial hypertension; PCSK9, proprotein convertase subtilisin/kexin type 9; VTE, venous thromboembolism. \*Cohorts shown are adults (age ≥ 18 years).*

### Cardiovascular and metabolic disease

Sample predictive tasks for future risk outcomes included coronary artery disease, heart failure, chronic kidney disease, and type 2 diabetes onset. For 1-year Future Risk tasks in adults (age ≥ 18) with no prior history of the outcome, the AUROC was 0.80 for coronary artery disease, 0.87 for heart failure, 0.81 for chronic kidney disease, and 0.80 for type 2 diabetes onset. Among Care Opportunity outcomes, GLP-1 receptor agonist eligibility was AUROC 0.86. These discrimination levels may support actionable stratification for preventive cardiology and metabolic care-gap closure.

### Oncology

Sample oncology outcomes included colorectal cancer (Screening, AUROC 0.73), prostate cancer (Screening, AUROC 0.75), multiple myeloma (Screening AUROC 0.84), and breast cancer (1-year Future Risk, AUROC 0.76).

### Musculoskeletal and functional outcomes

Sample outcomes ranged from the likelihood an individual will undergo a procedure to experience an acute event including: total knee replacement (1-year Future Risk, AUROC 0.82), total hip replacement (1-year Future Risk, AUROC 0.78), lumbar spine surgery (1-year Future Risk, AUROC 0.78), fragility fracture among patients with osteoporosis (1-year Future Risk, AUROC 0.76), and fall risk in adults older than 64 years (1-year Future Risk, AUROC 0.66). Procedure-oriented surgical outcomes generally showed higher discrimination than non-specific fall risk.

### Behavioral health and neurological outcomes

AUROCs for prediction of new-onset dementia (including Alzheimer’s disease) varied with age thresholds: 0.91 in the broadest adult cohort (age ≥ 18) to 0.80 in the oldest cohort (age ≥ 71). Additional neuropsychiatric outcomes examples include Parkinson’s Disease (Screening, AUROC 0.85), substance use disorder (Screening, AUROC 0.79), major depressive disorder in 65+ (Future Risk, 1 year, 0.73) and suicide-attempt (Future Risk, 1 year, AUROC 0.79). Screening outcomes for episodic mood disorders (e.g., bipolar disorder (AUROC 0.83) and major depressive disorder (0.72). Discrimination was strongest for progressive neurodegenerative conditions.

### Healthcare utilization

Sample outcomes included emergency-department utilization and unplanned hospitalization. Consistent with prior literature, AUROC values for acute utilization events were in the 0.64–0.66 range (emergency-department visits, general and age ≥ 65 cohorts), whereas all-cause hospitalization reached AUROC 0.76 in the general adult population and 0.68–0.74 in chronic-disease subpopulations (heart failure, CKD, diabetes, and cancer).

### Peri-procedural and treatment-effect outcomes

Treatment Effect predictions quantify the likelihood of a specific complication or outcome following a defined procedure, supporting individualized surgical risk assessment and post-acute care planning. Sample Treatment Effect predictive tasks presented were focused on surgical procedures for lower-extremity joint replacement, spinal fusion, coronary artery bypass grafting [CABG], surgical hip or femur fracture treatment [SHFFT], and major bowel procedures). AUROC across these tasks ranged from 0.63 to 0.85 (median 0.69). The strongest discrimination was observed for post-operative delirium following SHFFT (AUROC 0.85), 30-day all-cause readmission following SHFFT (0.79), and venous thromboembolism following total hip replacement (0.75); new chronic opioid use following spinal fusion also discriminated well (0.74).

Apart from SHFFT, 30-day all-cause readmission showed more modest discrimination across procedure types (AUROC 0.61–0.79 overall), consistent with the broader pattern that acute events are harder to discriminate than chronic progressive conditions.

### Cross-domain performance patterns

Several patterns emerged across the outcomes generated to date. First, chronic, progressive conditions (for example, chronic kidney disease [AUROC 0.81], heart failure [0.87], and dementia [0.84–0.91]) tended to show higher AUROC than acute or transient events (for example, emergency-department visits [0.64–0.66] and acute injuries [0.62–0.78]), consistent with prior literature.

### Benchmarking against published risk-prediction models

To place PhenOM performance in the context of tools already familiar to clinicians, we compared discrimination for six well-studied outcomes with the performance reported for established, guideline-referenced risk-prediction models (Table 3). For each of the six, PhenOM discrimination in the OM1 RWDC held-out data was comparable to or higher than the reported performance of the corresponding published models: Alzheimer’s disease and all-cause dementia (AUROC 0.92 and 0.91 vs. C-statistics of 0.49–0.84 for the CAIDE, ANU-ADRI, and UK Dementia Risk Scores), heart failure (0.87 vs. 0.71–0.85 for PCP-HF in external validation), myocardial infarction (0.85 vs. 0.67–0.81 for the Pooled Cohort Equations and SCORE2 in external validation, and 0.86–0.88 for QRISK3 in its validation cohort), stroke (0.84 vs. 0.68–0.74 for the revised Framingham Stroke Risk Profile), and breast cancer (0.76 vs. a pooled AUC of 0.60 for the Gail model/BCRAT and 0.70–0.71 for BOADICEA and IBIS). These comparisons are illustrative rather than head-to-head: the published instruments were derived and validated in different populations, over different time horizons (generally 10 years or longer, compared with 1 year for PhenOM), and using pre-specified predictor sets, and differences in case mix, outcome ascertainment, and prediction window preclude direct equivalence.

**Table 3.** Comparison of PhenOM Future Risk discrimination with published risk-prediction models for six representative outcomes.

| Outcome (Future Risk) | PhenOM AUROC (cohort) | Established model(s) | Reported discrimination (published) | Key reference(s) |
| --- | --- | --- | --- | --- |
| Alzheimer's disease | 0.92 (general population, age ≥18; 1 year) | CAIDE Dementia Risk Score; ANU-ADRI; UK Dementia Risk Score (DRS) | C-statistic* 0.77 (CAIDE, derivation); 0.49–0.60 (CAIDE, external, older adults) 0.64–0.74 (ANU-ADRI, external, Alzheimer's disease) 0.84 (UK DRS, age 60–79); 0.56 (UK DRS, age 80–95) | Kivipelto et al<sup>8</sup>; Anstey et al<sup>9,10</sup>; Walters et al<sup>11</sup> |
| Dementia (all-cause) | 0.91 (general population, age ≥18; 1 year) | CAIDE Dementia Risk Score; UK DRS | C-statistic 0.77 (CAIDE, derivation); 0.84 (UK DRS, age 60–79) | Kivipelto et al<sup>8</sup>; Walters et al<sup>11</sup> |
| Heart failure | 0.87 (general population, age ≥18; 1 year) | PCP-HF (Pooled Cohort equations to Prevent Heart Failure); Framingham HF; ARIC HF | C-statistic 0.71–0.85 (PCP-HF, external validation across race–sex groups) | Khan et al<sup>12</sup>; Kannel et al<sup>13</sup>; Agarwal et al<sup>14</sup>; Bavishi et al<sup>15</sup> |
| Myocardial infarction | 0.85 (general population, age ≥18; 1 year) | Pooled Cohort Equations (ASCVD, 2013 ACC/AHA); QRISK3; SCORE2 | C-statistic 0.67–0.72 (Pooled Cohort Equations, external validation) 0.67–0.81 (SCORE2, external validation) 0.86–0.88 (QRISK3, validation cohort) | Muntner et al<sup>16</sup>; Goff et al<sup>17</sup>; Hippisley-Cox et al<sup>18</sup>; SCORE2 Working Group<sup>19</sup> |
| Stroke | 0.84 (general population, age ≥18; 1 year) | Framingham Stroke Risk Profile (FSRP) | Revised FSRP C-statistic 0.68–0.74 (external validation) | Wolf et al<sup>20</sup>; Dufouil et al<sup>21</sup> |
| Breast cancer | 0.76 (general population, age ≥18; 1 year) | Gail model / BCRAT; Tyrer-Cuzick (IBIS); BOADICEA/CanRisk | AUC 0.60 (95% CI 0.58–0.62; Gail model, pooled) C-statistic 0.71 (IBIS), 0.70 (BOADICEA), 0.60 (BCRAT) (10-year, family-history-enriched cohort) | Gail et al<sup>22</sup>; Wang et al<sup>23</sup>; Tyrer et al<sup>24</sup>; Lee et al<sup>25</sup>; Terry et al<sup>26</sup> |

*PhenOM AUROC values are from the OM1 RWDC held-out data and reflect 1-year prediction windows. Comparisons are not head-to-head; published values reflect the cohorts, predictors, and time horizons (generally 10 years or longer) of the original studies. Published models were developed in narrower populations than the PhenOM cohorts (e.g., CAIDE in midlife adults, UK DRS in adults aged 60–95, Pooled Cohort Equations in adults aged 40–79, Gail model in women aged ≥35). CAIDE and UK DRS predict all-cause dementia rather than Alzheimer's disease specifically; the Pooled Cohort Equations,*

## Discussion

### Principal findings

The results presented here show that PhenOM can complete multi-domain predictive tasks for clinical outcomes at scale from a single real-world-data foundation-model platform. The thousands of predictive tasks completed and validated to date, covering more than 1,770 outcomes across 12 clinical domains (including healthcare utilization and mortality) and four outcome types, demonstrate this capability; the 177 tasks presented here are a sample that illustrates its breadth rather than a fixed or exhaustive catalog. Across all predictive tasks completed to date, 80% achieved AUROC of at least 0.70, and 30% achieved AUROC of at least 0.80. The fact that most predictive tasks achieved AUROC of at least 0.70 is broadly consistent with the literature on EHR-based prediction, where models are typically built for a single task; in contrast, the underlying approach allows additional predictive tasks for outcomes to be generated and validated as needed rather than requiring a separate model for each new question.

### Comparison with task-specific and EHR-embedded models

For the six outcomes in Table 3, PhenOM’s discrimination was comparable to or higher than the published performance of established task-specific risk scores. Widely deployed EHR-embedded tools such as Epic’s clinical decision support models have shown extensive site-to-site heterogeneity in external validation (I² ≥ 93%)^27^, which underscores the risk of evaluating models within narrow training environments; training and evaluation on the multi-source OM1 RWDC may reduce this source of generalizability risk, although this remains to be confirmed by external validation.

Our findings are also broadly concordant with APOLLO, also an EHR foundation model, on overlapping endpoints — for example, comparable discrimination for incident heart failure (0.87 here vs. 0.88) and myocardial infarction (0.79–0.85 vs. 0.82). Like APOLLO, PhenOM applies a single pretrained model across many predictions rather than task-specific models. The approaches are complementary: that model adds imaging and clinical-text modalities within one health system (7.2 million patients), whereas PhenOM is trained on a national, multi-source network of more than 400 million patients and is oriented toward generating an extensive array of validated, broadly representative, deployable outcomes usable in many contexts.^7^

### Implications for population health, patient management and drug development

Screening, Future Risk, Care Opportunity and Treatment Effect predictive tasks already have enormous applications across all healthcare sectors and tools like PhenOM will increase their availability and task specificity by orders of magnitude. Providers are using predictive tasks to reduce undiagnosed conditions, prioritize patient problem lists, close care gaps and counsel on the likely effects of treatment. Population health managers are similarly leveraging these tools for screening, care management prioritization, care gap closure and resource forecasting. Drug developers can support modeling of trial-eligible or endpoint-relevant cohorts or event predictions for observational and interventional research.

### Limitations

Several limitations should be acknowledged. First, all performance metrics reported here derive from the OM1 RWDC, a large and representative data set of U.S. patient journeys; to the extent a specific population is not well represented, results may not be applicable. Second, external validation has been limited to testing only a fraction of the available predicted outcomes in formal clinical validation studies (e.g., total knee arthroplasty, Fabry disease, abdominal aortic aneurysm, spinal fusion, etc.). Third, for some predictions, outcomes are defined using administrative and claims-derived code sets, which may undercount or misclassify some conditions relative to chart-review gold standards.

### Future directions

Expanding external validation partnerships with health systems and payers are planned to assess generalizability beyond the OM1 RWDC. As the underlying foundation-model architecture evolves, benchmarks will be updated and versioned to maintain a current, reproducible reference standard. In addition to a risk or likelihood score, PhenOM also surfaces the key predictive factors underlying each prediction, supporting clinical review, consistent with feature-attribution methods reported for contemporaneous EHR foundation models^7^.

## Conclusions

To date, PhenOM has been used to complete and validate thousands of predictive tasks covering more than 1,770 outcomes across four outcome types — Future Risk, Screening, Care Opportunity, and Treatment Effect — spanning major clinical domains including cardiovascular, metabolic, renal, oncologic, neuropsychiatric, musculoskeletal, surgical, and healthcare-utilization outcomes. The 177 tasks presented here are a representative sample illustrating this breadth, and the catalog continues to expand as new use cases and research questions arise.

Performance across the predictive tasks completed to date showed strong discrimination across a broad range of conditions, procedures and treatments. Taken together, these results offer a transparent, reproducible view of what a large-scale, real-world predictive AI platform can produce, and they invite comparison by independent groups applying the same outcome definitions to external data sources.

## Data Availability

Performance benchmarks reported in this manuscript are based on the OM1 Real-World Data Cloud. The terms of the data use agreements governing the RWDC prohibit third-party access to the data.

## Declarations

### Funding

This work was supported by OM1, Inc. There was no additional funding.

### Competing interests

All authors are employees or officers of OM1, Inc. OM1 is a for-profit healthcare AI company, and PhenOM is a commercial product of OM1.

### Ethics approval

The protocol describing use of the RWDC for research on patient clinical characteristics and outcomes was reviewed by the Advarra Institutional Review Board and determined to be exempt. All data were de-identified, and informed consent was not required.

### Code availability

PhenOM is a proprietary model; its code and model weights are not publicly available.

### Author contributions

Conceptualization: [CB, JB, RG]. Methodology: [MM, KV, JB, CB]. Formal analysis: [JB, MM, KV, ML]. Writing – original draft: [ML, RG]. Writing – review and editing: all authors. MM and KV contributed equally. All authors approved the final manuscript.

### Use of AI-assisted tools

AI-based writing tools were used to assist with editing portions of this manuscript. The authors reviewed and edited all content and take full responsibility for the final manuscript.

## Notes

### Author Declarations

Advarra Institutional Review Board. OM1, Inc., Use of Real-World Data to Understand Patient Characteristics, Risk Factors, Treatment Patterns, and Outcomes across Care Settings and Condition Areas (Pro00074434). Determined to be exempt 16 Oct 2023.

